# The accuracy and repeatability of OpenEvidence on complex medical subspecialty scenarios: a pilot study

**DOI:** 10.64898/2025.11.29.25341091

**Authors:** Jawahar Jagarapu, Kikelomo Babata, Surya Chamarthi, Robert Hoyt

## Abstract

OpenEvidence is a popular artificial intelligence (AI) based medical search engine that generates evidence-based answers. It includes a quick search engine method (OE) that takes only seconds to respond, along with a limited number of references. In mid-2025, the platform introduced “Deep Consult” (DC), which takes several minutes to respond and provides more comprehensive answers with additional references. OpenEvidence scored 100% on USMLE-type multiple-choice questions, but it has not been tested on more complex medical scenarios. We tested the OE and DC models using questions primarily derived from medical specialty board exams, specifically, the MedXpertQA dataset. In a prior published study, this dataset was evaluated with eleven large language models (LLMs), and the results indicated poor accuracy (14-46%) for all LLMs. We evaluated the performance of OpenEvidence on a sample of the MedXpertQA dataset, comprising 100 medical subspecialty scenarios and using two independent evaluators.

The highest accuracy for DC was 41%, and for OE, 34%. Repeatability testing revealed an evaluator concordance rate of 77% for OE and 72% for DC.

## Introduction

The practice of medicine mandates extensive knowledge, critical thinking, and experience. Clinicians, however, often encounter situations where they are uncertain about the diagnosis or treatment plan. They also frequently need to expand the differential diagnoses that are to be considered. This is when access to high-quality, up-to-date information is paramount. Textbooks are often outdated, and a literature search may be slow and nonproductive. This is why online resources such as UpToDate, DynaMed, and Epocrates are popular. The complexity of navigating UpToDate’s vast content and its cost have presented challenges. In mid-2025, UpToDate launched an artificial intelligence (AI)-enabled version that allows searches to utilize natural language processing. [1]

Another recent addition to the medical armamentarium is OpenEvidence. [2] OpenEvidence was created by Daniel Nadler, PhD, in 2023 as part of the Mayo Clinic Platform Accelerate program. The results are evidence-based, and content agreements have been established with the Journal of the American Medical Association (JAMA) and the New England Journal of Medicine (NEJM) for sharing full-text content. Many of the OE and DC answers include images or tables from one of these resources. OE has also partnered with Elsevier’s ClinicalKey AI to supply medical content. OpenEvidence is web-based but also available on iOS and Android, making information retrieval convenient. OpenEvidence has two search features. The first one is a quick search or Consult; from now on, we will refer to it as “OE.” In OE, a clinical question is entered into the main search window in natural language, which then generates a response with citations after the text. The references are hyperlinked to online abstracts. Following the references is the ability to ask follow-up questions. The second feature is called “Deep Consult,” referred to as “DC.” Deep Consult was introduced in mid-2025 and delivers comprehensive reports with many more references, but the reports take longer to generate. It is touted to deliver “PhD level” expertise, and according to one source, DC is an agentic AI system, more capable than OE, but with higher computational requirements. [5] No studies have evaluated its accuracy to date.

In addition to its medical search engine, OE provides various other features. OE can create patient handouts, generate tables, calculate risk scores, create multiple-choice questions based on external reviews, and draft prior authorization letters. [2–3] OpenEvidence achieved a score of 100% on the United States Medical Licensing Examination (USMLE) in 2025. It’s free for physicians in the United States who possess an NPI number, as well as for some locations overseas. It is very popular and is said to have 400,000 users as of July 2025. [2] According to Dr. Nadler, OE is “an ensemble of specialized models rather than a single large model,“ all trained exclusively on peer-reviewed medical literature without connection to the public internet.” This suggests that OE uses retrieval augmented generation (RAG). [4]

Studies evaluating OE’s accuracy and completeness have been limited. Hurt et al. studied five common medical problems in primary care: hypertension, hyperlipidemia, diabetes mellitus type 2, depression, and obesity using OE. The impact on clinical decision-making was minimal, despite the high scores for clarity, relevance, support, and satisfaction. In other words, it reinforced plans rather than modifying them. [6] Low et al. compared the performance of OE, ChatRWD, and three LLMs on 50 open-ended clinical scenarios. Nine physicians rated the results for accuracy and relevance. ChatRWD achieved 58%, followed by 24% for OE and 2-10% for the LLMs. [7] Hajj et al. compared ChatGPT-4o and OpenEvidence clinical decision support for structural heart disease. They submitted 15 questions to each model related to transcatheter tricuspid valve repair or replacement. They found that ChatGPT-4o produced more reliable answers, as judged by subject matter experts. [8]

Despite its widespread use, several concerns have been raised. Patel et al. wrote about the shortcomings of OpenEvidence, including a lack of transparency, the process by which articles are curated or excluded, the timeliness of the information, and its clinical relevance. [9] Furthermore, there is a knowledge gap regarding OpenEvidence’s performance in complex subspecialty clinical scenarios. OE and DC should be tested on more challenging medical datasets to evaluate their reasoning on questions that are not found online. Subspecialty questions are more complex than general primary care questions and, as a result, require more in-depth knowledge to answer. It is recognized that standard medical board exam questions, such as those found in the USMLE, are no longer adequate because they don’t comprehensively test medical reasoning. [10]

In 2025, Zuo et al. published an article on the creation of the MedXpertQA dataset. This dataset includes 4,460 questions obtained from the USMLE, COMLEX-USA, and 17 US medical specialty boards, covering 11 body systems. The questions were filtered using probabilistic semantic similarity to remove highly similar questions. The questions were also slightly modified to reduce the likelihood that LLMs encountered them during training. The possible answers were expanded to ten choices, making decision-making more difficult. Lastly, the questions were reviewed by physicians for accuracy and validity, although their specialty status is unknown. The dataset is divided into “Text” for text evaluation and “MM” for multimodal evaluation. The text category was further divided into reasoning and understanding scenarios.

Twelve large language models were tested, and on the reasoning data subset, the highest accuracy score was 46% by GPT-o1, and the lowest was 14% by Qwen2.5-32B. [11] The accuracy results for text-reasoning questions across all models studied are available in the supplement.

### Objective

The objective is to evaluate the accuracy of OE and DC models using the MedXpertQA dataset. Results will be compared with the LLMs reported in Zuo et al.’s article. [11] Furthermore, OE and DC lack repeatability data, so two researchers will test the same medical scenarios to ensure concordance. One hundred randomly selected text-based reasoning scenarios will serve as the basis for this pilot study. We hypothesized that the accuracy of OE and DC would be higher than that of the publicly available LLMs when evaluated on the MedXpertQA dataset.

## Methods

### Dataset

The MedXpertQA dataset was downloaded from a data science platform in the CSV file format. [12] The dataset was filtered to include only text-related questions and only the reasoning category. This resulted in 2447 multiple-choice questions with 10 possible answer choices (A-J). The average length of the total number of characters in the questions was 1,173, with a range from 268 to 4,527. Python 3.12 was used to categorize the questions by body system and length: short (<940 characters), medium (940-1363 characters), and long (>1363 characters). The distribution was as follows: short (25%), medium (50%), and long (25%). [13] The questions were also divided into diagnosis (49.5%), treatment (33.9%), and basic science (16.6%). The percentage of questions listed by body system is listed in the Supplement.

Python 3.12 was also used to stratify and randomize the questions. The data were sorted by stratum, and the number of samples per stratum was calculated. The Python libraries involved were pandas, numpy, sklearn.model_selection, ast, and argparse. For example, “Basic Science_Respiratory_Medium.” Systematic random sampling was performed within each stratum, and no duplicate questions were found. Python selected 100 scenarios for a pilot test. The initial unfiltered dataset, the pilot dataset of 100 scenarios, the final datasheet with links to all answers, and the corresponding Python code for stratified sampling are available on GitHub.

1. [14] A sample question is included in the supplement, with hyperlinks to the answers provided by the OE and DC methods.

### Testing Procedure

An application programming interface (API) exists, but access is limited and not publicly available. Therefore, two authors (JJ and RH) manually submitted the same multiple-choice questions (MCQs) to OE and the DC to test accuracy and repeatability. The questions were submitted as is with no attempt at prompt engineering.

### OE Query

After pasting scenarios into OpenEvidence, searches involved two phases: “analyzing query” followed by “finalizing report.” The results were available in seconds and were always accompanied by several references. The time spent analyzing the query and finalizing the report was recorded, as were the number of references and the query’s uniform resource locator (URL).

### DC Query

After pasting scenarios into Deep Consult, searches involved several phases: “reasoning,” then “searching medical literature,” and finally “preparing final report.” A reasoning phase may reappear occasionally after reviewing the medical literature. Following the initial query, DC asks 3-5 questions to obtain more details. Because we had no further clinical input, we responded with, “No further information is available.” DC searches automatically published the time it took to reason and research the literature. That time was automatically recorded and displayed on the OpenEvidence platform, along with the number of references and the query URL. Finalizing the report generally took as long as the initial phases, but this time was not recorded.

### Data Collection

All the data were recorded in a Google AppSheet application, which has a user-friendly interface and a database backend. [15] For each independent evaluator and each search method, we collected the following data elements: the selected answer from the multiple-choice options, the time taken for the model to process the query, the number of references per output, and the public links for the search results. The AppSheet data was downloaded to a CSV file after the study was complete for further analysis.

## Evaluation Metrics

1. The overall accuracy of OE and DC for each evaluator
2. The repeatability between evaluators for OE and DC
3. The accuracy by body system per evaluator
4. The number of references and the range for both OE and DC per evaluator
5. The response times for both OE and DC per evaluator
6. The correlation of accuracy with body system, question length, medical task, and the number of references
7. Answers: When both OE and DC are correct and incorrect. When OE is correct and DC is incorrect, and when OE is incorrect and DC is correct per evaluator

## Statistics

We conducted all the statistical analyses and visualizations using Python version 3.12 and its statistical libraries, NumPy, pandas, matplotlib, and seaborn. Descriptive statistics were performed. We evaluated the accuracy of OE and DC methods against the ground truth for each method and rater, as well as the overall accuracy for each method by body system, question length, and medical task. The chi-square test compared the accuracy of OE and DC methods, while the kappa statistic assessed inter-rater concordance. A Pearson correlation evaluated the relationship between reference count and accuracy. The Mann-Whitney U test compared OE and DC based on reference counts and response times. One-way ANOVA calculated the significance of medical tasks and question length on accuracy across groups.

## Results

### Accuracy by method and evaluator

The accuracy of OE and DC is displayed in **Figure 1**. The OE accuracy for RH and JJ is 28% and 34%, respectively, with an overall average of 31%. The DC accuracy for RH and JJ is 41% and 38%, respectively, with an overall average of 39.5%. While the average accuracy of DC is higher than that of OE, a chi-square analysis showed no statistically significant difference (p-value = 0.09). There was a 6% difference between the two reviewers (RH and JJ) for OE and a 3% difference for DC. The instances where OE was correct, but DC was incorrect for evaluators RH and JJ are 3% and 6%, respectively. Similarly, in the cases where DC was correct and OE was incorrect, the percentages for RH and JJ are 16% and 10%, respectively. For both evaluators, only 25% of the time OE and DC were correct together, and 75% of the time they were discordant. In some instances, for both evaluators, the model output showed a different answer that was not among the available multiple-choice options. We listed them as answer “K.”

**Figure 1:**
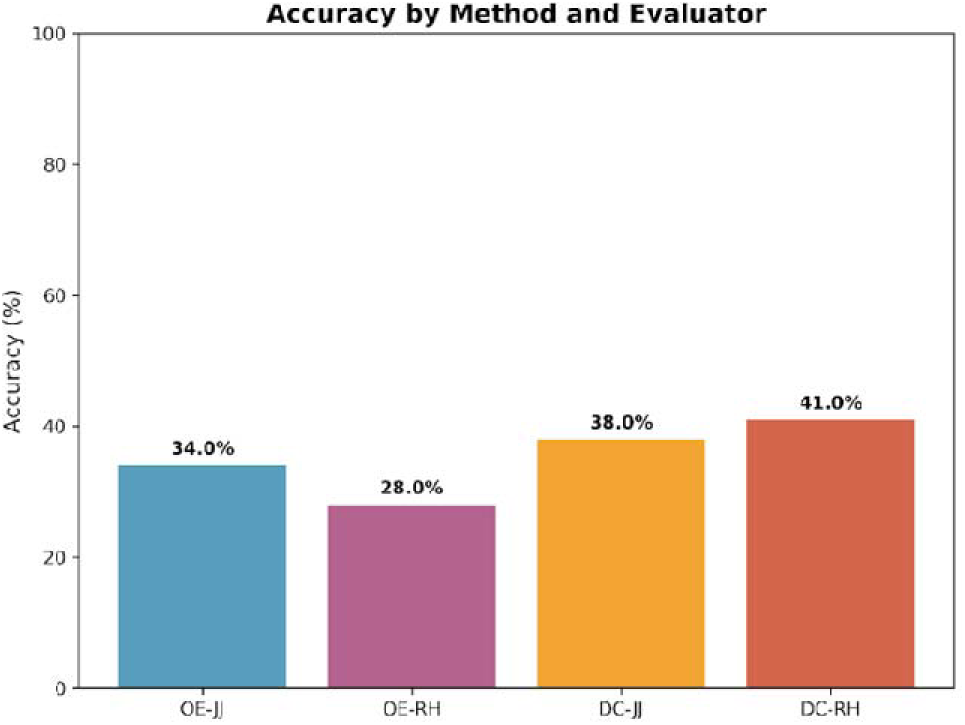
Overall accuracy by method and evaluator. For the OE method, OE-JJ (34%) has the highest accuracy, and for the DC method, DC-RH (41%) has the highest accuracy.

For the OE method, the K answers were 2% for both evaluators; for the DC method, they were 4% and 6% for RH and JJ, respectively.

### Repeatability

Figure 2 shows the concordance between intra-and inter-methods for both evaluators, regardless of accuracy. The concordance for the OE and DC methods between RH and JJ was 77% and 72%, respectively. The concordance between methods across evaluators was 60%. Cohen’s kappa measure (κ) of overall agreement between evaluators for both OE and DC methods was 0.74 and 0.69, respectively, indicating substantial agreement.

**Figure 2:**
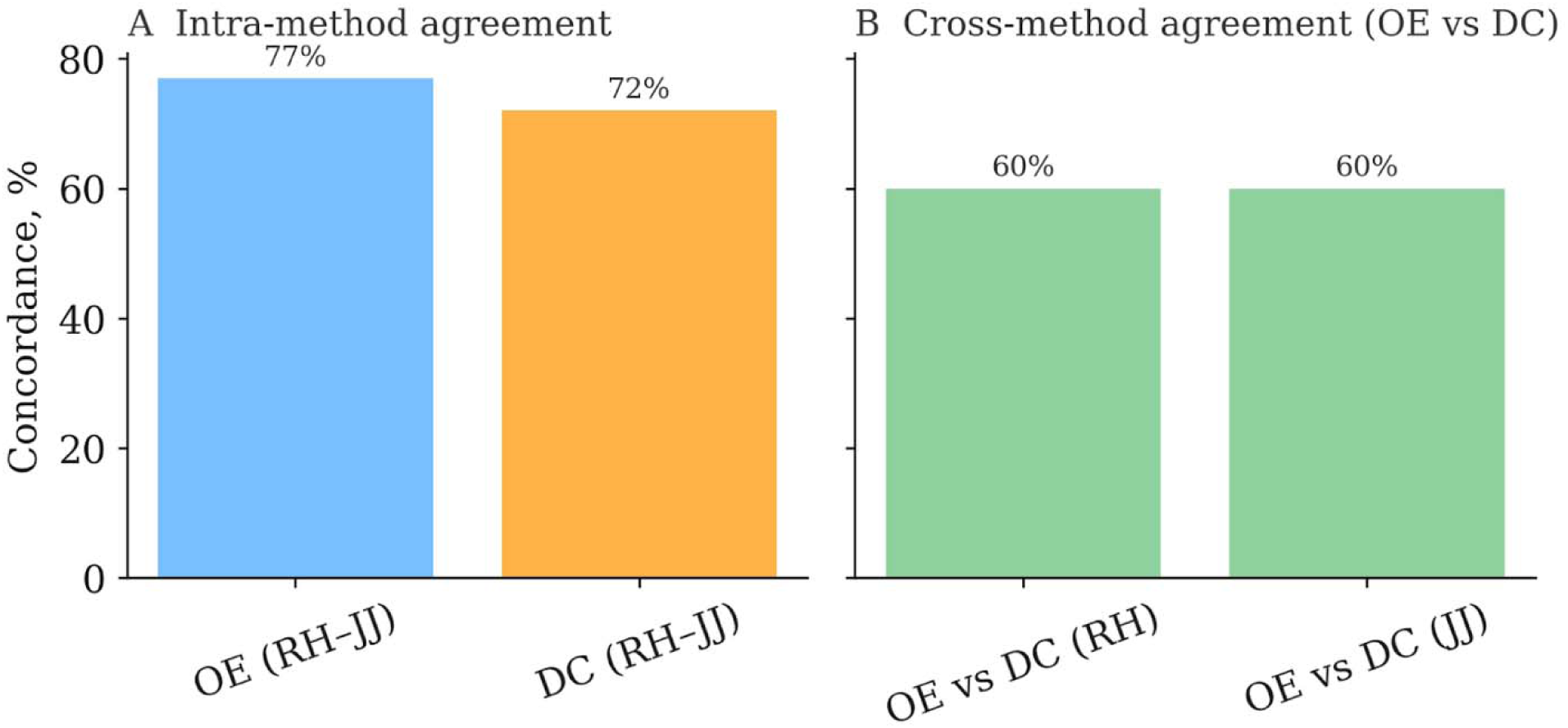
Concordance between methods and evaluators. Figure 2A: Concordance between evaluators for OE and DC; Figure 2B: Concordance between the methods for each evaluator, RH and JJ. Cohen’s kappa (κ) value for intra-method agreement for OE (0.74) and for DC (0.69). **Cohen’s Kappa value interpretation:** ≤ 0 - no agreement, 0.01–0.20 as none to slight, 0.21–0.40 as fair, 0.41–0.60 as moderate, 0.61–0.80 as substantial, and 0.81–1.00 as almost perfect agreement.

### Accuracy by Body Systems

**Table 1** reports on the accuracy of OE and DC for the body system and evaluator. The highest and lowest average accuracies for OE by body system were muscular (42.8%) and skeletal (21.9%), respectively, and for DC, digestive (55%) and respiratory (30%), respectively. The question counts were low for most body systems, so these results should be interpreted with caution.

**Table 1:** OE and DC accuracy by body system, and evaluator (Top 7 body systems)

| Body system | Count | OE-JJ | OE-RH | OE-Average | DC-JJ | DC-RH | DC-Average |
| --- | --- | --- | --- | --- | --- | --- | --- |
| Nervous | 17 | 23.5% | 35.2% | 29.4% | 33.3% | 33.3% | 33.3% |
| Skeletal | 16 | 25% | 18.7% | 21.9% | 31.2% | 56.2% | 43.7% |
| Cardiovascular | 12 | 33.3% | 50% | 41.7% | 33.3% | 33.3% | 33.3% |
| Respiratory | 10 | 20% | 30% | 25% | 30% | 30% | 30% |
| Endocrine | 10 | 30% | 30% | 30% | 40% | 40% | 40% |
| Digestive | 9 | 22.2% | 44.4% | 33.3% | 55% | 55% | 55% |
| Muscular | 7 | 42.8% | 42.8% | 42.8% | 42.8% | 57.1% | 50% |

### References and Accuracy

For OE, the number of references for evaluator RH ranged from 1 to 12 with a mean of 5.2 and SD of 2.2, and for JJ, the number ranged from 1 to 11 with a mean of 5.16 and SD of 2.2. For DC, for evaluator RH, the number of references ranged from 0 to 84 with a mean of 35.7 and SD of 16.5, and for JJ, the number ranged from 6 to 96 with a mean of 33.8 and SD of 15. Overall, for OE, the average is 5.2 ± 2.2 references, and for DC, the average is 34.7 ± 15.8 references.

Figure 3 shows the reference counts per method and evaluator and average references per method. The distribution for references deviated from normality, so we used non-parametric testing. OE used significantly fewer references than DC, with a significant Mann–Whitney U test (U = 349.5, p = 6.18e-65; n = 200 per group), indicating strong stochastic dominance of lower reference counts in OE.

**Figure 3:**
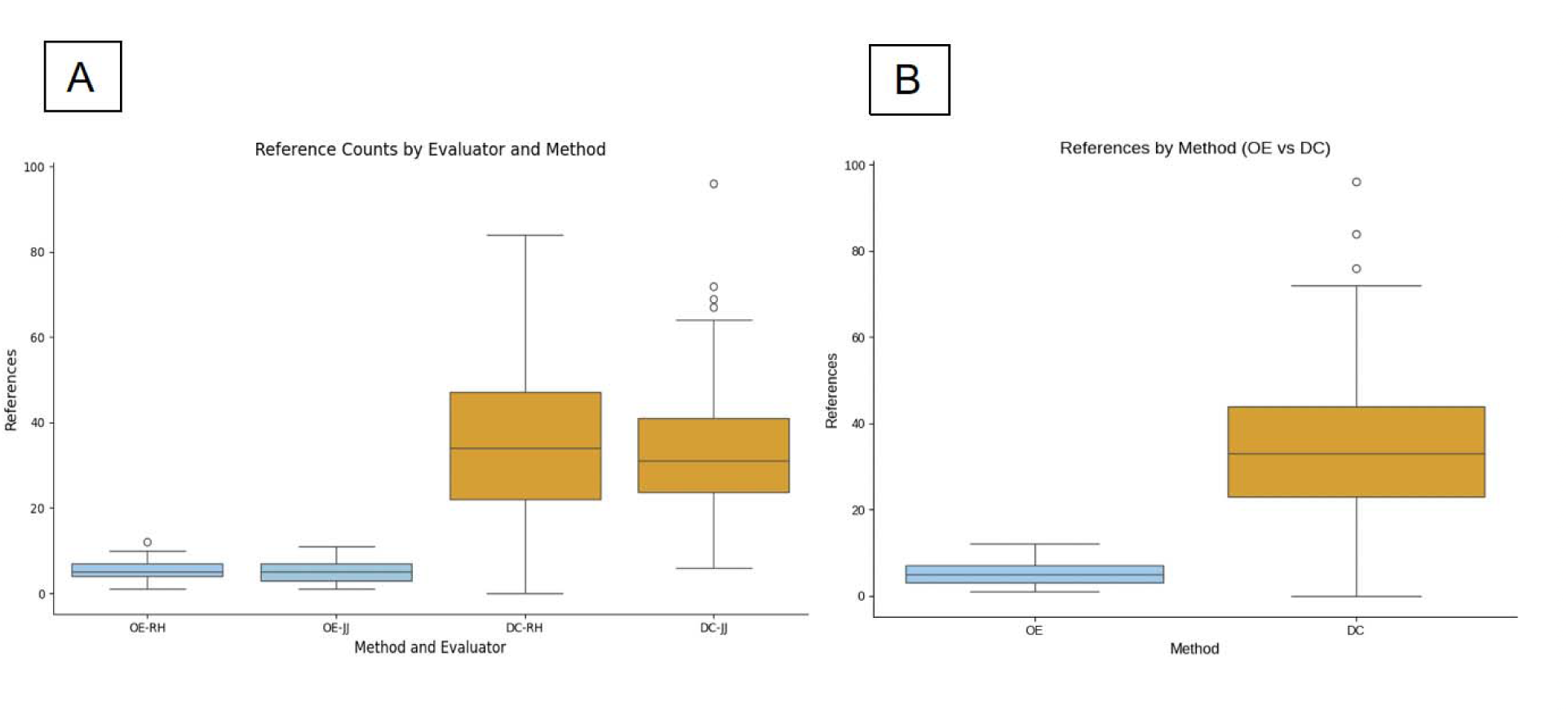
References per method and evaluator. Figure 3A: Reference Counts per Method and Evaluator. Each box shows the interquartile range (IQR), with the horizontal line marking the median; whiskers denote the minimum and maximum values. OE-RH (n=100): mean 5.2, SD 2.2, median 5 (IQR 4–7), range 1–12; OE-JJ (n=100): mean 5.2, SD 2.3, median 5 (IQR 3–7), range 1–11; DC-RH (n=100): mean 35.7, SD 16.5, median 34 (IQR 22–47.3), range 0–84; DC-JJ (n=100): mean 33.8, SD 15, median 31 (IQR 23.8–41), range 6–96. Figure 3B: Overall References by Method. Box plots display the distribution of reference counts for the OE and DC methods. Each box represents the interquartile range (IQR: 25th–75th percentile); the horizontal line indicates the median, and whiskers extend to the minimum and maximum observed values. For OE references (n = 200): mean 5.19, SD 2.2, median 5 (IQR 3–7), range 1–12. For DC references (n = 200): mean 34.7, SD 15.8, median 33 (IQR 23–44), range 0–96.

### Correlation between Reference Count and Accuracy

Figure 4 displays the distribution of accurate OE and DC responses in relation to the number of displayed references. (OE, n = 62 accurate out of 200 responses; DC, n = 79 accurate out of 200 responses). The Pearson correlation coefficient is 0.11 for OE and-0.01 for DC and is statistically insignificant. Overall, there is no significant correlation between reference count and accuracy (Pearson correlation: r = 0.068, p-value = 0.177). There is no correlation between the number of references and accuracy for both methods.

**Figure 4.**
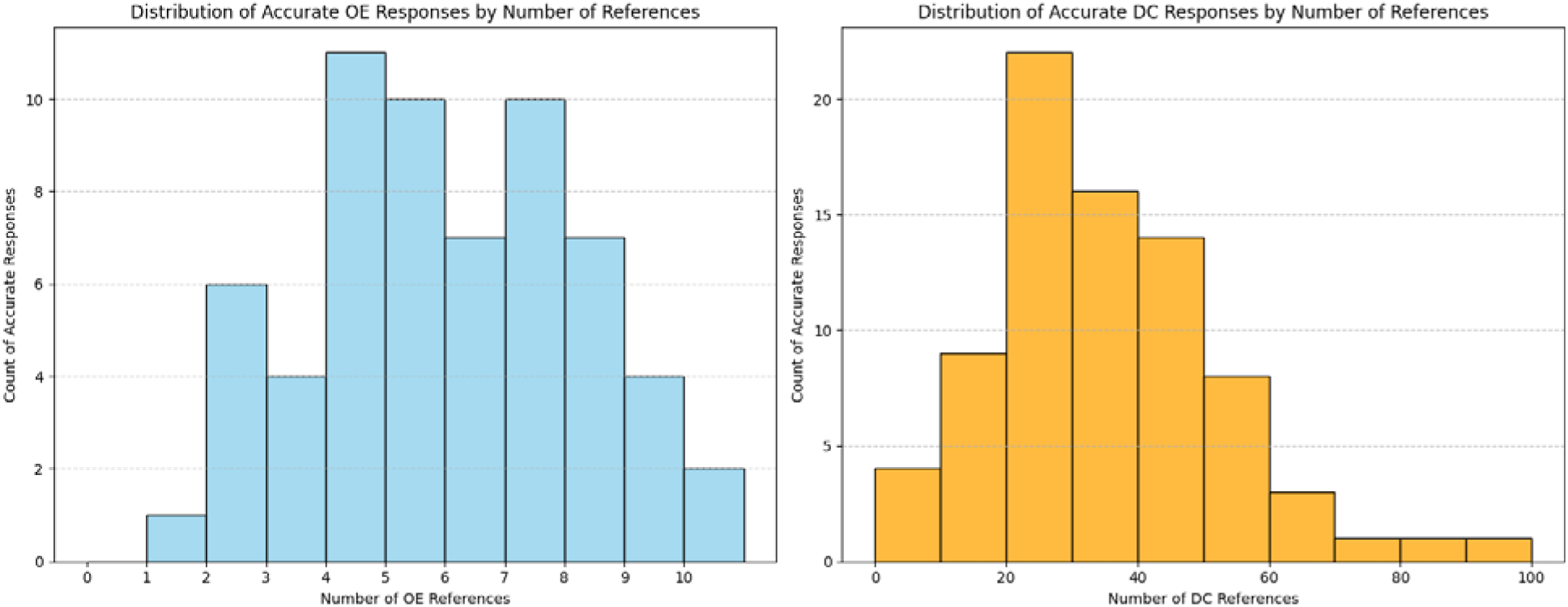
References versus accuracy for OE and DC. Histograms show the distribution of the number of references for overall accurate responses for OE (left) and DC (right) methods.

### Model Response Times and Accuracy

Figure 5 displays the response times for OE and DC methods. For OE, the mean response times for RH and JJ were 15 and 11 seconds, respectively. For DC, the mean response times for RH and JJ were 271 and 247 seconds, respectively. The overall average time for OE was 13 seconds, and for DC, 259 seconds. OE response times were significantly faster than DC. The time difference was statistically significant, as assessed by the Mann–Whitney U test (U = 0.0, p = 1.28e-67; n = 200 per group). Figure 6 shows the distribution of accurate responses versus time for OE and DC methods. The Pearson correlation values for accuracy versus time are not statistically significant (OE, r = 0.15, p = 0.145), (DC, r = 0.02, p = 0.84.) 0.145).

**Figure 5:**
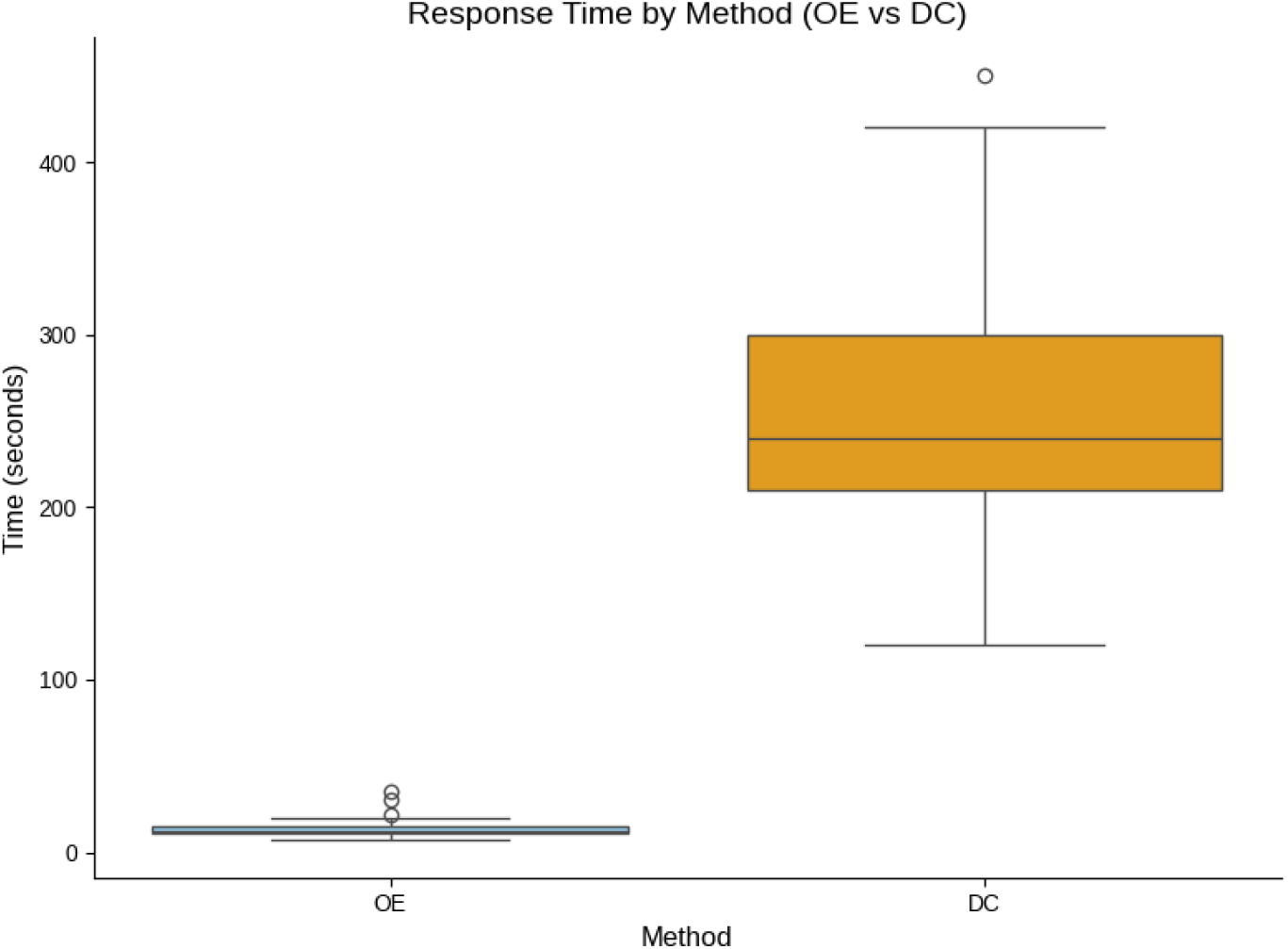
Response Times by Method (rounded to the nearest second). Each box represents the IQR (25th–75th percentile), with the line indicating the median. OE (n=200): mean 13 s, SD 6 s, median 13 s (IQR 10–15 s), range 4–57 s. DC (n=200): mean 259 s, SD 83 s, median 240 s (IQR 180–300 s), range 120–600 s

**Figure 6:**
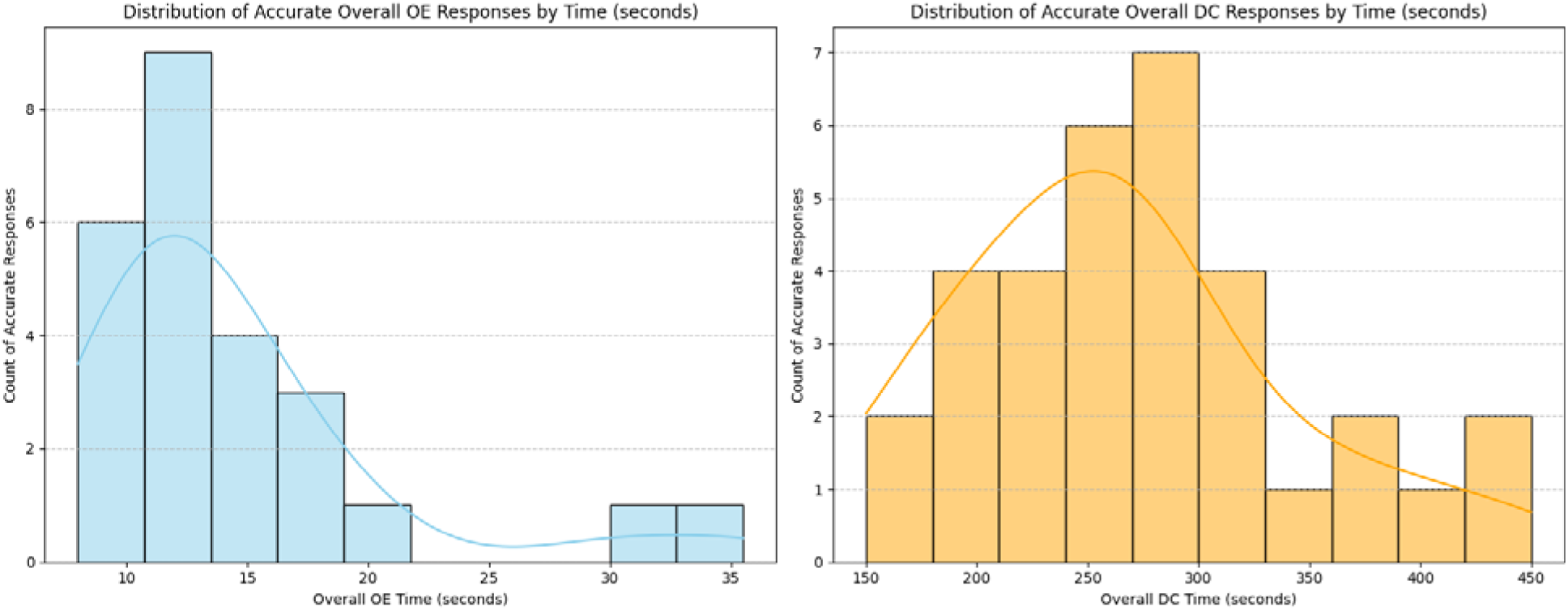
Response Time versus Accuracy distribution for OE and DC methods. Histograms show the distribution of accurate overall OE and DC responses against the time displayed (in seconds).

### Accuracy and Repeatability by Medical Task

**Basic Science:** These tasks show the lowest mean accuracies for both OE and DC responses by both raters (OE-JJ: 23.5%, OE-RH: 23.5%, OE-Average 23.5%, DC-JJ: 29.4%, DC-RH: 35.3%, DC-Average 32.4%). Basic science tasks, on the other hand, have the highest OE repeatability (82.4%), which means that raters are consistent even if their accuracy is low. The DC repeatability for basic science is the lowest (64.7%).

**Diagnosis:** Accuracy is slightly higher than basic science for DC responses (DC-JJ: 36.0%, DC-RH: 38.0%, DC-Average: 37%) but remains relatively low for OE responses (OE-JJ: 28.0%, OE-RH: 22.0%, OE-Average: 25%). Both the OE and DC repeatability are moderate at 72.0% and 70.0%, respectively.

**Treatment:** Treatment tasks exhibit the highest mean accuracies among the three categories (OE-JJ: 48.5%, OE-RH: 39.4%, OE-Average: 43.9%). DC-JJ: 45.5%, DC-RH: 48.5%, 48.5%, DC-Average: 47%. Repeatability is also high for treatment tasks, with OE at 81.8% and DC at 78.8%.

Figure 7 shows the overall accuracy of methods and medical tasks. In summary, ‘Treatment’ tasks appear to have the highest accuracy and repeatability, while ‘Basic Science’ tasks show the lowest accuracy. One-way ANOVA tests were conducted to determine if the specific medical task (e.g., Basic Science, Treatment, Diagnosis) influenced the accuracy of responses for both the OE and DC methods. The results indicated no statistically significant influence, suggesting that the type of medical task does not meaningfully affect the overall response accuracy for either the OE or DC methods within this dataset.

**Figure 7:**
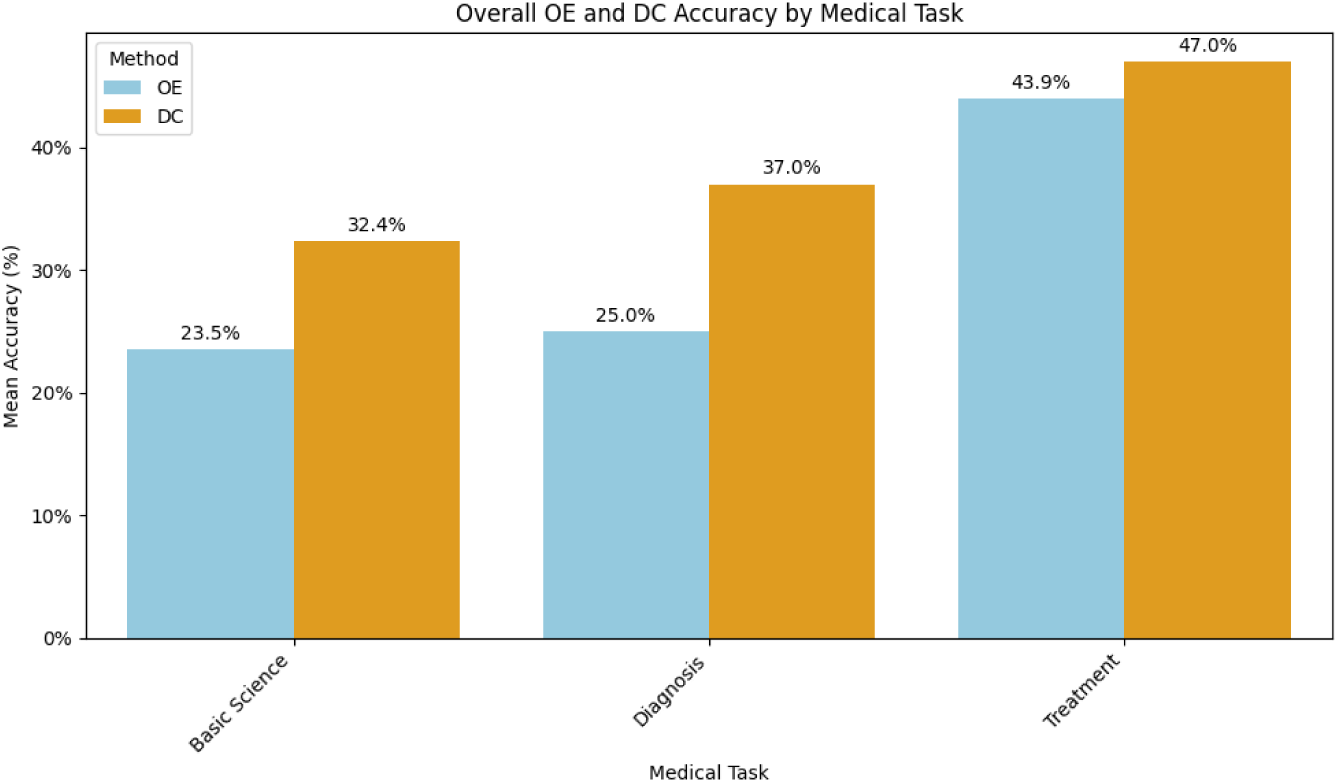
Accuracy by Method and Medical task. The overall mean accuracy was determined by the medical tasks of basic science (OE-23.5%, DC-32.4%), diagnosis (OE-25%, DC-37%), and treatment (OE-43.9%, DC-47%). One-way ANOVA tests between three groups showed no statistical significance for each method. OE method (F statistic = 2.28, p-value = 0.107) and DC method (F statistic = 0.72, p-value = 0.489)

### Accuracy and Repeatability by Question Length

**Short:** The accuracy for ‘Short’ questions was low for both raters for the OE method (OE-JJ: 23%, OE-RH: 29%, OE-Average: 26.5%), whereas as for DC method the accuracy is relatively high (DC-JJ: 35%, DC-RH: 47%, DC-Average: 41.2%)

**Medium:** The accuracy for ‘Medium’ questions for both raters and methods are as detailed (OE-JJ: 36%, OE-RH: 26%, OE-Average: 31.1%, DC-JJ: 39%, DC-RH: 41%, DC-Average: 40.6%).

Compared to ‘Short’ questions, the overall OE average accuracy increased for medium questions, while the DC average accuracy decreased.

**Long:** The accuracy for ‘Long’ questions for both raters and methods are as detailed (OE-JJ: 36.7%, OE-RH: 30%, OE-Average: 33.3%, DC-JJ: 36.7%, DC-RH: 36.7%, DC-Average: 36.7%). Compared to ‘Medium’ questions, the overall OE average accuracy increased for ‘Long’ questions, while the DC average accuracy decreased.

For OE, the repeatability between raters is relatively high and consistent across all question lengths:’Short’ (82%),’Medium’ (75%), and’Long’ (76%). For DC, the repeatability is more variable, with the highest for’Long’ (83%), the lowest for’Short’ (47%) and moderate for’Medium’ (73%) question length.

Figure 8 displays overall accuracy by question length and method. In summary, for the OE method, there is a trend towards increased accuracy with longer question length, whereas for the DC method, we observed a downward trend in accuracy with longer question length. One-way ANOVA tests were conducted to determine if the ‘Question length’ (e.g., Short, Medium, Long) influenced the accuracy of responses for both the OE and DC methods. The results indicated no statistically significant effect, suggesting that ‘Question length’ does not affect overall response accuracy for either the OE or DC methods in this dataset.

**Figure 8:**
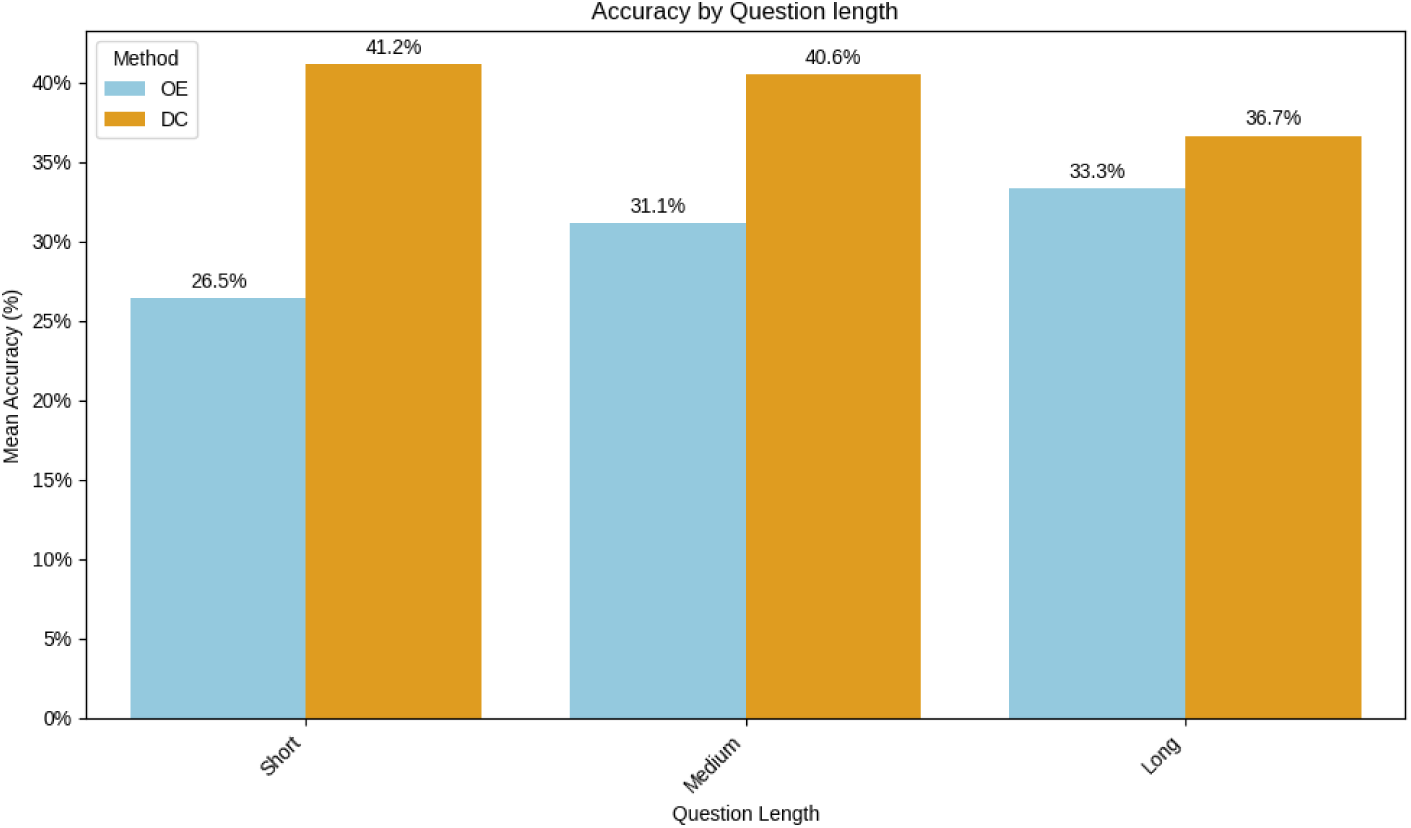
Accuracy by Question length and Method. One-way ANOVA tests between three groups showed no statistical significance between question length and accuracy for the OE method (F-statistic = 0.14, p-value = 0.87) and the DC method (F-statistic = 0.08, p-value = 0.92).

## Discussion

### Overall Accuracy and Benchmarking

This study demonstrates that OE and DC do poorly at answering complex subspecialty multiple-choice questions. The results for OE are similar to several LLMs tested by Zuo et al. [11] Deep Consult performed better than OE but was no better than the leading LLM. The highest accuracy scores achieved (41% for DC and 34% for OE) were similar to what has been published for other frontier models utilizing the MedXpertQA dataset but lower than expected given Open Evidence’s reported progression from ∼90% to 100% on USMLE-style questions. [16] The outcome underscores the substantial challenge posed by expert questions at the subspecialty level, along with the importance of having expert oversight in complicated clinical scenarios, since OE has been shown to have limitations in specialty settings. [8] It’s crucial also to consider another essential performance metric, repeatability, which we defined as whether the same question given to an LLM generated the same answer. Prior LLM studies have described this quality as “LLM consistency” [17] or being “stable.” [18] We defined repeatability as the same-answer rate under fixed settings in a short time window (also termed “same-answer stability“) and reported it as the proportion of times with identical final answers when the question was repeated (two times) by two evaluators.

The evaluator-level repeatability was 77% for OE and 72% for DC. The observed evaluator-level repeatability metrics further illuminate the challenges and limitations of using LLMs in clinical decision-making. It should be noted that repeatability does not even translate into accuracy. [17] These repeatability rates were lower than those reported by human experts. For instance, pathologists interpreting breast biopsies for invasive breast cancer demonstrate overall agreement rates of 89-92% intra-and inter-observer. OpenEvidence’s repeatability was, however, higher than what has been found for more challenging breast lesions, such as atypia (43-53%). [19] In other clinical contexts, higher interrater agreement has been observed on the same question within short intervals; for example, κ≈0.89–0.92 for end-of-life medical orders. However, even in these cases, 8% showed at least one discordant order across interviews, demonstrating that perfect agreement remains uncommon even in high-stakes clinical settings. [20]

Though repeatability was substantial between evaluators, low accuracy is concerning, given emerging evidence that authors are already trialing and recommending OpenEvidence and other large language models as adjuncts to clinical reasoning. [6] The probabilistic nature of LLMs, which generates outputs from probability curves instead of deterministic answers, contributes to this response variability and helps explain why similar questions lead to different results in the OE and DC models. LLMs have also been shown to have significant metacognitive deficiencies and are often unaware of their limitations; experts have suggested not relying solely on performance on board examinations as a metric of evaluation. [21] In the legal domain, similar accuracy and repeatability concerns have been published, showing that leading models can return unstable answers to the same question. Their findings reinforce the need to report LLM stability metrics alongside accuracy for end users. [18] Recent specialty evaluations comparing OE with frontier models reach a similar cautionary conclusion: neither tool should be used without expert oversight, and outputs are best employed as adjuncts to and not replacements for clinical judgment. [8] Because expert oversight alone cannot guarantee reliable outputs, systematic human validation is critical to ensuring consistency and trust in model performance

### Comparative Performance of OE vs. DC methods

The study results indicate that the accuracy of the model output increased from the OE method to the DC method for both raters, ranging from 4% to 13% (relative improvements of 11.7% to 46.4%). There were no prior studies that compared the two methods of OpenEvidence in the setting of text-based, complex medical subspecialty board-style questions. Another study evaluated the standard versus deep research approach to medical reasoning on 1000 human-curated clinical questions (MedBrowseComp dataset) and found a 34–75% relative improvement in accuracy in the OpenAI o3 and Gemini 2.5 Pro models, respectively. While there was a significant relative improvement in the deep consult query compared to the standard query, the overall accuracy was still below 40% for the models. [22] Although we did not provide additional clarifying questions, when compared to OE, the DC search required a significantly longer time to retrieve answers, accompanied by a substantially higher number of references, which was reflected in the increased overall accuracy (though not statistically significant). The underlying mechanism for DC search likely involves an agentic search framework compared to OE search, where the model asks for additional clarifying questions and then displays a series of reasoning steps while performing the search, like the agentic mode operations seen in the deep research option of publicly available commercial LLMs like ChatGPT, Perplexity, and others.

It seems reasonable that if DC were given additional information to answer its follow-up questions, its performance would significantly improve; however, a recent study by Stanford suggests otherwise. They introduced script concordance testing (SCT), a validated medical assessment tool designed to evaluate clinical reasoning. SCTs measure how the addition of new information can alter diagnostic and treatment decisions. They developed a publicly available benchmark comprising 750 SCT questions drawn from 10 international medical datasets across multiple specialties and clinical domains. They compared ten frontier LLMs with opinions by medical students, residents, and faculty. The models tested matched or exceeded medical students’ performance on many SCT tests, but their results were lower than those of senior residents and expert clinicians. The data suggest that LLM and DC accuracy may not improve significantly with the addition of further information. [23]

### Model Response Times and References

Though the DC method performed better than OE in accuracy, it came with a significant time tradeoff. The median DC response time was almost 18 times longer than the median OE response time (240 vs. 13 seconds). In addition, the median number of references is six times higher in the DC search compared to the OE search (33 vs 5), which significantly increases the time to verify the sources. This result prompts a question about when to use quick search and deep consult methods in an LLM-based evidence search engine. The authors believe that for straightforward clinical scenarios that require a quick point-of-care knowledge retrieval (e.g., guideline-based knowledge, established medical practices, recent evidence lookup), the OE method might be suitable. In contrast, for complex scenarios that require multi-step reasoning and detailed analysis (e.g., rare clinical presentations or topics that need an evidence update), the DC might be an appropriate method. “OE” mode may suit *clinician-in-a-hurry* use cases, while “DC” may align better with *research and evidence synthesis*. For either method, clinicians should verify the model output by checking reference sources and their interpretation, given the low accuracy.

### Human Validation Workflow and Reliability

Our research illustrates the value of using multiple raters to validate LLM outputs in evaluation studies. The accuracy and repeatability of a model lay the foundation for trusting its future use, especially in complex medical settings where reasoning is required. If the model proposes a different approach for diagnosis or treatment when tested in two instances, that could result in low trust among the users for its future use as a point-of-care clinical decision support tool. OE and DC outputs from our study findings show moderate to substantial repeatability between the two raters. When considering the deployment of more LLM-based clinical decision support models, it’s crucial to validate their output against ground truth by different raters. The inter-rater agreement validation should be a part of human verification when deploying these models for clinical decision support in the future. We will need standardized validation protocols involving dual or multiple raters for evaluating model outputs in clinical settings. We also observed that the model generated a different answer that was not included in the proposed multiple-choice options infrequently in both OE and DC methods. Neither model stated, “I don’t know the answer.”

### Nature of the MedXpertQA Dataset and Its Influence

The MedXpertQA dataset consisted of multiple-choice questions of varying lengths and complexities, as well as medical tasks (basic science, diagnosis, and treatment). We did not see a significant change in the accuracy performance of the model between short and long question lengths. However, the accuracy performance based on the medical task was the lowest for basic science tasks and the highest for treatment-related questions. The authors note that the ground truth for the answers was established by the publishers of the MedXpertQA dataset, which was used as a comparison for accuracy and accuracy testing in this study. We chose not to validate the ground truth of the dataset. However, in the context of complex clinical reasoning scenarios, there may often be two or even three plausible answers or approaches to the diagnosis or treatment in a real-world setting. This problem poses a significant challenge in evaluating LLM outputs based on a single ground truth answer in medical reasoning.

## Limitations

As a pilot study, it is likely it is underpowered for subgroup analyses. In this initial pilot phase, we did not conduct error analyses because it would have required obtaining subject-matter expert opinions across multiple subspecialties. We limited our evaluation of OE/DC to a single subspecialty dataset, recognizing the existence of alternative approaches. Errors are likely, and experts may disagree on the right diagnosis or treatment. Deep Consult’s extensive literature search could yield fresh insights, given that the board review questions could be several years old. Focusing on JAMA and NEJM resources might impact the accuracy of results. We did not analyze all references for pertinence and the presence of hallucinations. The study results suggest that while OE/DC may be better suited for general medical inquiries, its suitability for subspecialty questions is unclear. Larger studies are necessary to confirm this finding. Our evaluation was constrained by limited access to the OpenEvidence API, preventing a full dataset assessment; thus, we relied solely on the pilot dataset with manual data entry. Furthermore, we lack information regarding technical details of OE/DC models, temperature settings, and the agentic approach, the model’s retrieval-augmented generation (RAG) architecture, specifically whether it utilizes vector or graph-based methods.

### Future Recommendations

The use of LLMs in clinical care carries significant risks. Our findings underscore several essential requirements for the secure deployment of LLMs in clinical reasoning and decision support. First, standardized human validation of LLM outputs remains essential given the performance limitations observed. Second, developers of medical LLM tools must conduct rigorous testing across varying levels of clinical complexity and specialties and provide transparent reporting of these results. Clinicians need clear information about model limitations, particularly the discrepancy between performance on basic medical knowledge questions versus complex clinical scenarios. As we have shown, models that perform excellently on medical student-level questions may show significant accuracy limitations when faced with the complexity of real-world clinical decision-making. Finally, we recommend establishing dual validation and transparent benchmarking as standard practice for evaluating medical LLMs to ensure reliable performance metrics and support informed decision-making about their clinical applicability. We recommend that OE/DC be tested on open-ended questions and complex scenarios using a resource it likely did not train on, such as BMJ case reports. [24]

## Conclusions

OpenEvidence, performed with low accuracy on this sample MedXpertQA dataset, like what has been found with publicly available LLMs. The repeatability rates for the dataset are lower than expected, which is concerning given clinicians have started to utilize these tools at the point of care for clinical decision support. These findings emphasize that there must be expert oversight when using these tools in clinical care. We need further large studies to validate their accuracy and repeatability on various complex subspecialty medical scenarios, case reports, and other medical resources.

## Abbreviations

AI: Artificial Intelligence
OE: OpenEvidence Quick consult method
DC: OpenEvidence Deep consult method
LLM: Large Language Model
CSV: Comma Separated Value
S: Seconds
RAG: Retrieval augmented generation
URL: Uniform Resource Locator

## Data Availability

GitHub Repository. Available from: https://github.com/rehoyt/MedXpertQA

