## Supplement for "The accuracy and repeatability of OpenEvidence on complex medical subspecialty scenarios: a pilot study"

1. Model accuracy (derived from [7])

| **Model** | **Accuracy** |
| --- | --- |
| GPT-o1 | 46.2 |
| Expert | 41.7 |
| DeepSeek R1 | 36.8 |
| GPT-o3 mini | 36.6 |
| DeepSeek V3 | 23.9 |
| LLaMA-3.3-70B | 23.8 |
| QVQ-72B-Preview | 22 |
| LLaMA-3-70B-UltraMedical | 20 |
| QwQ-32B-Preview | 18.7 |
| Qwen2.5-72B | 18.5 |
| Claude-3.5-Haiku | 16.7 |
| Qwen2.5-32B | 14 |

2. Questions by Body System

| **Specialty** | **Percent (%)** |
| --- | --- |
| Nervous System | 15.4 |
| Cardiovascular | 14.1 |
| Skeletal | 12.8 |
| Digestive | 11.6 |
| Respiratory | 8.9 |
| Endocrine | 8.1 |
| Reproductive | 8.1 |
| Muscular | 7.1 |
| Urinary | 5.2 |
| Other/NA | 4.2 |
| Lymphatic | 2.9 |
| Integumentary | 1.7 |

3. Sample question (Original text question #53)

A 64-year-old male arrives at the emergency department via ambulance with symptoms that began two hours ago, including difficulty speaking, right facial numbness, and weakness in his right arm and leg. His medical history includes type 2 diabetes mellitus, hypertension, hyperlipidemia, and a 25 pack-year smoking history. He takes insulin, metformin, lisinopril, and simvastatin. His vital signs show: temperature 37.0°C (98.6°F), heart rate 80/min, respiratory rate 16/min, and blood pressure 140/85 mmHg. Physical examination reveals an alert patient who follows commands but cannot correctly identify the current month or his age. He exhibits right facial droop, right-sided limb drift, and diminished sensation on the right side, without eye deviation. What should be the immediate next step in managing this patient? Answer Choices: (A) Carotid duplex ultrasound (B) Computerized tomography (CT) of the head (C) Intravenous (IV) tissue plasminogen activator (tPA) (D) Aspirin (E) Fingerstick glucose level (F) Electrocardiogram (ECG) (G) Neurological consultation (H) Magnetic resonance imaging (MRI) of the brain (I) Echocardiogram (J) Complete blood count with coagulation profile.

4. Link to answer by OE by evaluator JJ: <https://www.openevidence.com/ask/34a25815-356f-4504-a652-5f3cf0df9f41>

            Link to answer by DC by evaluator JJ:

<https://www.openevidence.com/ask/049d92d3-969b-4edf-880e-8a507e30466a>
